# Use of Powered Air-Purifying Respirator(PAPR) by healthcare workers for preventing highly infectious viral diseases -a systematic review of evidence

**DOI:** 10.1101/2020.07.14.20153288

**Authors:** Ana Licina, Andrew Silvers, Rhonda L Stuart

**Author notes:** **Corresponding Author** Dr Ana Licina, Mailing Address: 145 Studley Road Heidelberg Victoria 3084. **Disclosure Statement:** Authors have no significant financial or non-financial disclosures to make.

## Abstract

**Background:** Healthcare workers (HCWs) are at particular risk during pandemics and epidemics of highly virulent diseases with significant morbidity and case fatality rate. These diseases include Severe Acute Respiratory Syndrome Coronaviruses, SARS-CoV-1 and SARS-CoV-2, Middle Eastern Respiratory Syndrome (MERS) and Ebola. With the current (SARS-CoV-2) global pandemic, it is critical to delineate appropriate contextual respiratory protection for HCWs. The aim of this systematic review was to evaluate the effect of Powered Air Purifying Respirators (PAPR’s) as part of respiratory protection versus another device (egN95/FFP2) on HCW infection rates and contamination.

**Methods:** Our primary outcomes included HCW infection rates with SARS-CoV-2, SARS-CoV-1, Ebola or MERS when utilizing PAPR. We included randomized controlled trials, non-randomized controlled trials, and observational studies. We searched the following databases: MEDLINE, EMBASE, and Cochrane Library (Cochrane Database of Systematic Reviews and CENTRAL). Two reviewers independently screened all citations, full-text articles, and abstracted data. Due to clinical and methodological heterogeneity, we did not conduct a meta-analysis. Where applicable, we constructed Evidence Profile (EP) tables for each individual outcome. Confidence in cumulative evidence for each outcome was classified according to the GRADE system.

**Results:** We identified 689 studies during literature searches. We included 10 full text studies. A narrative synthesis was provided. Two on-field studies reported no difference in the rates of healthcare workers performing airway procedures during care of critical patients with SARS-CoV-2. A single simulation trial reported a lower level of cross-contamination of participants using PAPR compared to alternative respiratory protection. There is moderate quality evidence that PAPR use is associated with greater heat tolerance but lower scores for mobility and communication ability. We identified a trend toward greater self-reported wearer comfort with PAPR technology in low quality observational simulation studies.

**Conclusion:** Field observational studies do not indicate a difference in healthcare worker infection utilizing PAPR devices versus other compliant respiratory equipment. Greater heat tolerance accompanied by lower scores of mobility and audibility in PAPR were identified. Further pragmatic studies are needed in order to delineate actual effectiveness and provider satisfaction with PAPR technology.

Please note: Protocol for this review was prospectively registered with the International Register of Systematic Reviews identification number CRD42020184724.

## Background

High infectivity combined with high case fatality rate during the COVID-19 pandemic have placed an emphasis on healthcare worker (HCW) protection both from a personal as well as a societal perspective. Several other outbreaks of virulent highly infectious diseases have occurred in recent decades including the Ebola crisis in 2014-2016, Middle East respiratory syndrome coronavirus (MERS-CoV) and Severe Acute Respiratory Syndrome (SARS, due to SARS-CoV-1) epidemic (1) (2). Teasing out the true infection risk in HCW group is difficult. This is due to the high rates of community infection, HCW travel and under-reporting of non-HCW populations and the lack of phylogentic viral analysis.. Personal protective equipment (PPE) and infection control guidelines from the WHO are based on the assumption that the primary mechanism of transmission of SARS-CoV-2 is direct and indirect droplet spread as well as fomite transmission (3). The WHO advises that airborne transmission can occur, but only when aerosol-generating procedures (AGPs) are performed in patients infected with SARS-CoV-2 (4). Aerosol Generating Procedures result in generation of small aerosolized particles through disruption of surface tension of the alveolar lining (5). Aerosolized particle clouds can travel up to 8 meters (6). A detailed, list of AGPs is provided in *Table 1* (7). The degree of airborne spread in the Coronavirus group is contentious (8, 9). Recently, stability of SARS-Cov-2 and SARS-Cov-1 was evaluated under different experimental conditions (10). SARS-CoV-2 and SARS-CoV-1 remained viable in aerosols throughout the three hour duration of the experiment with a reduction in infectious titer (10). However the clinical relevance of this experimental model has been questioned (11). Establishing with certainty whether SARS-CoV-2 is infectious through airborne transmission may be methodologically challenging.

**Table 1.**
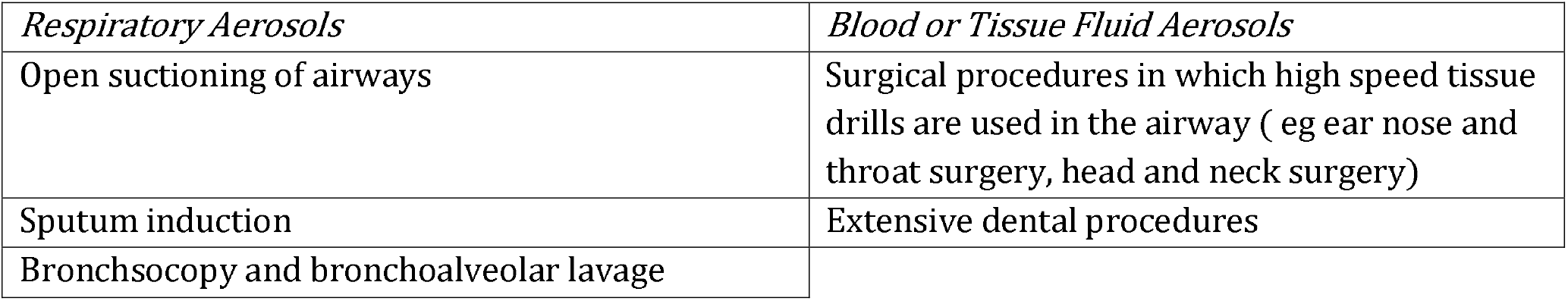
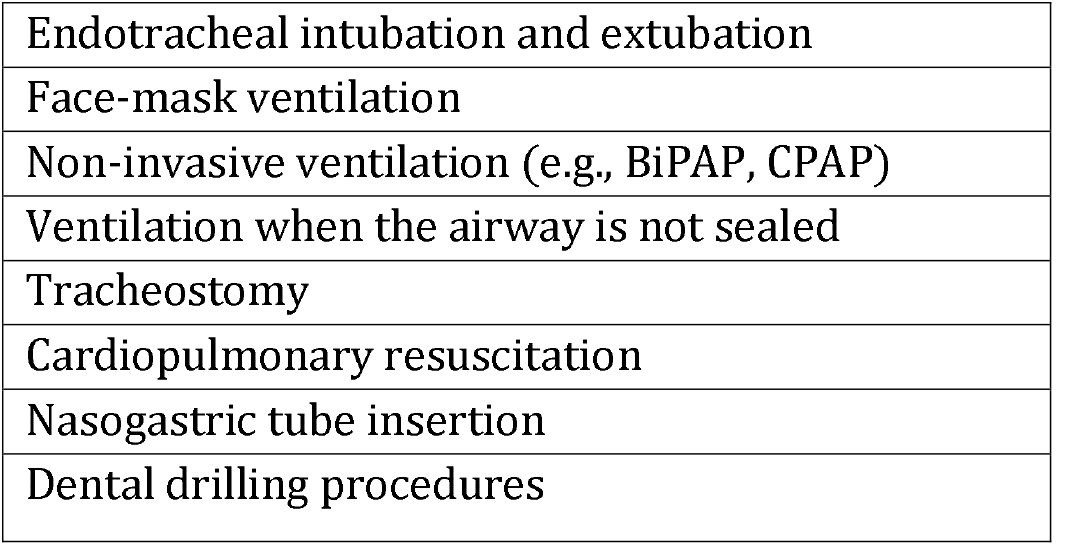
List of Aerosol Generating Procedures (AGPs) Abbreviations: BiPAP (Bilevel positive ventilation pressure), CPAP (Continuous positive airway pressure);

| *Respiratory Aerosols* | *Blood or Tissue Fluid Aerosols* |
| --- | --- |
| Open suctioning of airways | Surgical procedures in which high speed tissue drills are used in the airway ( eg ear nose and throat surgery, head and neck surgery) |
| Sputum induction | Extensive dental procedures |
| Bronchoscopy and bronchoalveolar lavage |  |

|  |
| --- |
| Endotracheal intubation and extubation |
| Face-mask ventilation |
| Non-invasive ventilation (e.g., BiPAP, CPAP) |
| Ventilation when the airway is not sealed |
| Tracheostomy |
| Cardiopulmonary resuscitation |
| Nasogastric tube insertion |
| Dental drilling procedures |

In this review, we consider the implication for HCWs of Ebola in addition to the coronaviruses. Ebola virus can be transmitted by direct contact with blood, bodily fluids, or skin of Ebola patients or individuals who have died of the disease. Development of Ebola disease results in a high case fatality rate, as high as 50%. Recommendations for respiratory protective equipment are therefore similar(12).

### Description of the Devices

Two major international testing and classification bodies of respiratory protection include the National Institute for Occupational Safety and Health (NIOSH) and European Norms (EN). Air-purifying particulate respirators function by removing aerosols (droplets and solid particles) from the air through the use of filters, cartridges or canisters. Air-purifying respirators fall into one of four different classifications (*Table 2*): 1. Filtering Facepiece Respirator (FFR), 2. Elastomeric Half facepiece Respirator, 3. Elastomeric Full Facepiece Respirator, and 4. Powered Air-Purifying Respirator (PAPR). The two major testing institutions (NIOSH and EN) employ different test protocols for evaluation of air-purifying particulate respirators as well as having different nomenclatures (*Table 2*). In the US, respiratory filtration levels are determined according to Occupational Health and Safety Administration (OSHA) standard 29 CFR 1910.134 “Respiratory Protection” (13). Meanwhile, the EN requires 94 and 99% efficiencies for FFRs, class P2 (FFP2) and class P3 (FFP3), respectively (14). In Europe respirators are tested against the relevant European Standard and are approved to the PPE Directive 89/686/EEC or the replacement PPE Regulation (EU)2016/425 (15).

**Table 2.**
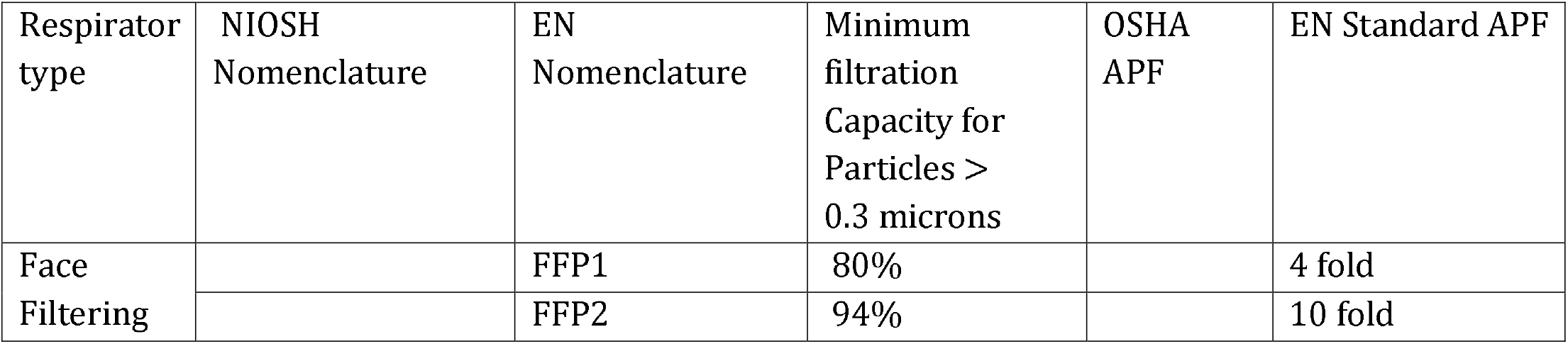
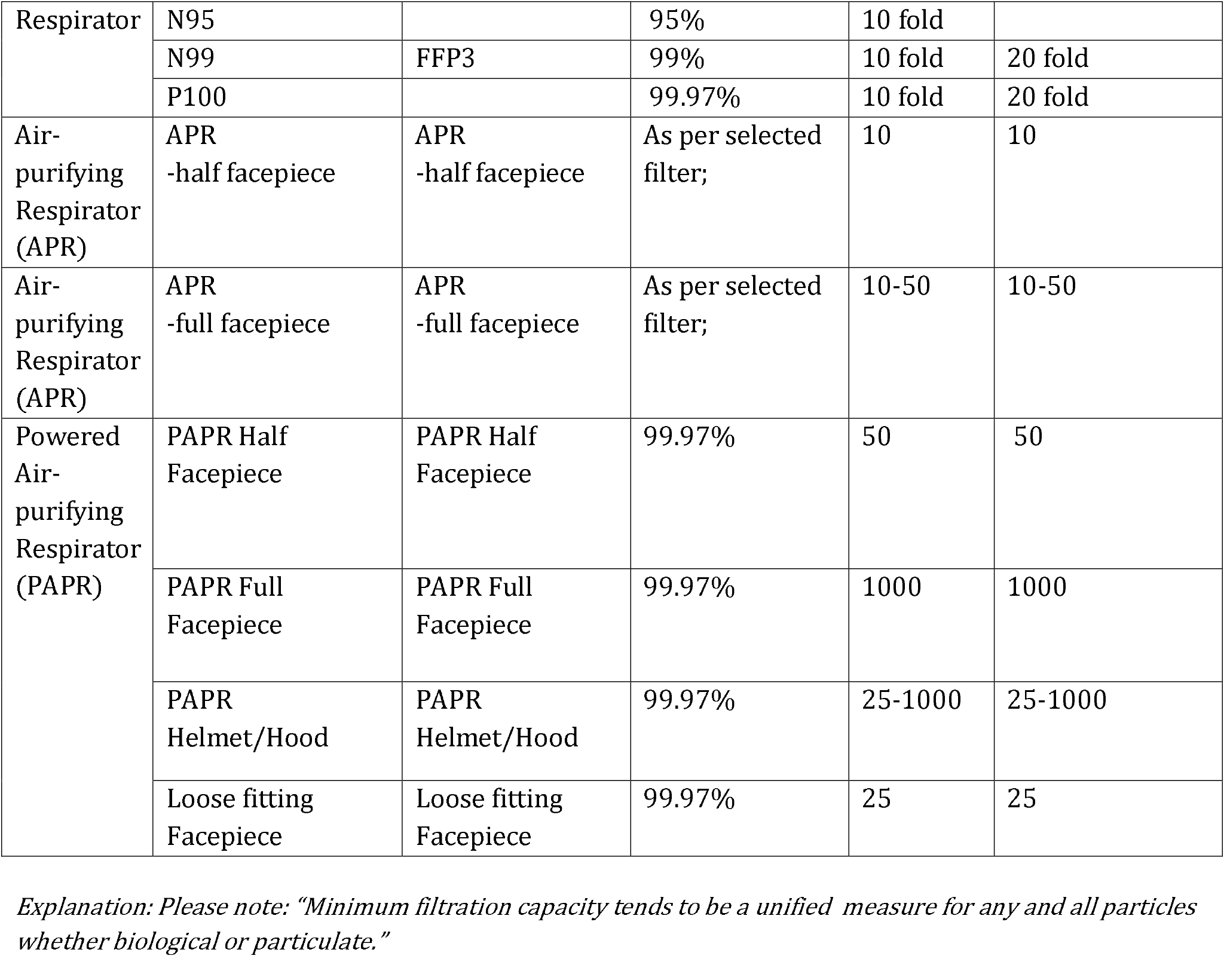
Filtering Face Piece, Air-Purifying Respirator(APR) and Powered Air-Purifying Respirator (PAPR) Classification according to NIOSH/EN (National Institute for Occupational Safety and Health (NIOSH) and European Norms (EN) with stated Assigned Protection Factor (APF)

The assigned protection factor (APF) of a respirator denotes the level of protection that the respirator is expected to provide to users who are properly fitted and trained. The APF is the ratio of pollutant outside the device (environment) to that inside the device (inhaled component). For example, an APF of 10*” means that a user could expect to inhale no more than one tenth of the airborne contaminant present”*. Airborne level protection includes: helmets, covers, hoods; FFP3 or FFP2/N95 masks, googles or face shields (if no helmets).

PAPRs can be described as respirators that protect the user by filtering out contaminants in the air and use a battery-operated blower to provide the user with clean air through a tight-fitting respirator, a loose-fitting hood, or a helmet. There is a wide heterogeneity of the available PAPR devices. Traditional PAPRs used in healthcare settings have a full-face piece and loose-fitting hoods, attached to waist mounted belt batteries. PAPRs use high-efficiency particulate air (HEPA) filters and provide a higher level of protection than disposable respirators. High-efficiency particulate air (HEPA) filters have a similar filtration as P100 (i.e., they filter at least 99.97% of particles 0.3 μm in diameter and are oil proof (9). PAPR’s are considered more protective in terms of the level of respiratory protection due to the higher efficiency of their filtration pieces as well as the maintenance of outward positive pressure. PAPRs are specified for high-hazard procedures because they can offer assigned APFs ranging from 25 to 1,000, which reduce the risk more than the protection factors provided by N95 respirators. The improved protection is largely provided by the positive pressure in the head covering or facepiece (*Table 2)*. The hoods of PAPRs can provide splash protection and some degree of eye protection (14, 16). If HCWs are provided with sufficient comfortable and well-fitting respiratory protection, it is likely that compliance with preventive programs will be increased (17).

### How the intervention might work

In the first instance, relevant individual institutions need to safeguard an HCW respiratory compliance program. Appropriate choice of the level of respiratory protection needs to be made within this program.

There is significant heterogeneity of international recommendations with regards to appropriate respiratory protection for HCWs when performing AGPs in suspected or confirmed COVID-19 patients is notable. European Centre for Disease Prevention and Control (ECDC) prevention and Centre for Disease Control (CDC) USA recommend use of an least N95/FFP2 and higher level of protection (18) (7); Public Health England recommends FFP3 level respiratory protection in addition to standard PPE (19); The Communicable Disease Network Australia (CDNA) recommends FFP2/N95 mask; With regards to the use of PAPR CDNA recommends that if a healthcare worker is required to remain in the room for longer periods of time (greater than one hour), the use of PAPR may be considered for additional comfort and visibility (20).

### Why it is important to do this review

Evidence guided practice for the respiratory component of personal protective equipment is limited. With the high rate of HCW infection during the SARS-COV-1 epidemic in Toronto, PAPR use became embedded in respiratory protocols (21) (22). Limited information exists for use of one type of facial protection (e.g. FFP3) over another (e.g. FFP2/N95). High filtration pieces appear to have a protective advantage in laboratory settings (23). However, this does not translate to firm findings of greater healthcare workers protection in field studies (24). Increased layers and technical challenges of personal protective equipment can lead to increased complexity of patient care (25). During outbreaks such as the current global pandemic, early recommendations are often based on precautionary principles. It is uncertain what level of respiratory protection is required routinely for aerosol generating procedures (AGP’s) in highly infectious viral diseases as evidenced by heterogenous international recommendations. In 2005, Yassi et al identified recommended level of respiratory protection as a critical gap in societal understanding of viral pandemic management (26). There are known logistical advantages and disadvantages to PAPR technology (see *Table 4*).

**Table 3.** International recommendations of respiratory component of PPE for protection of HCWs performing AGPs in suspected or conformed COVID-19 patients Abbreviations: Face Filtering Piece (FFP), Personal Protective Equipment (PPE), Healthcare workers (HCW), Aerosol Generated Procedures (AGP);

| International Governing Body/Institution | Face Filtering Piece (FFP) (in addition to other PPE) | Powered Air-Purifying Respirator (PAPR) (in addition to other PPE) |
| --- | --- | --- |
| European Centre for Disease (ECDC) | FFP2/FFP3 | Use of PAPR not considered |
| Centers for Disease Control and Prevention (CDC) | At least N95 | Use of PAPR not considered |
| Public Health of England | FFP3 | Use of PAPR not considered |
| The Communicable Disease Network Australia (CDNA) | FFP/N95 | Consider the use of PAPR if remaining in room with patient with suspected/confirmed COVID-19 positive patient longer than one hour; |

**Table 4.**
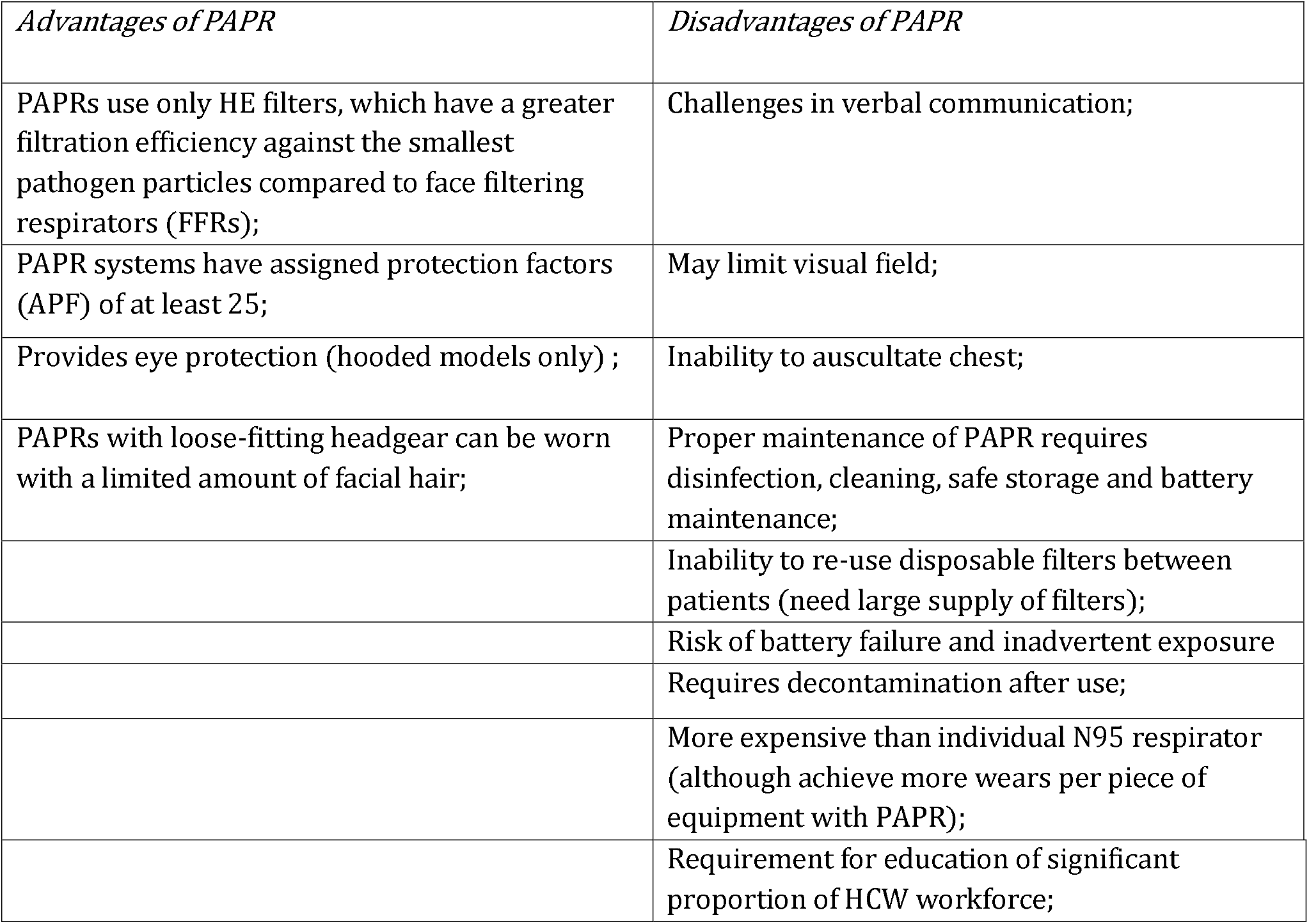
Logistical Advantages and Disadvantages of PAPR, adapted from Wong et al (27)

We aim to summarize and critically appraise current evidence of the effectiveness of PAPR for preventing nosocomial infection in health care staff exposed to respiratory/body fluids contaminated with highly infectious viral diseases such as SARS-CoV-2, SARS-CoV-1, MERS and Ebola. In particular, we will try and address current questions identified from the COVID-19 epidemic, that include to what effect PAPR as part of respiratory protection versus another (e.g.N95/P2) has on HCW infection rates and contamination.

## Methods

Our findings have been reported according to the standards for the Preferred Reporting Items for Systematic Reviews and Meta-Analysis (28). Protocol for this review was prospectively registered with the International Register of Systematic Reviews identification number CRD42020184724.

### Eligibility criteria Types of studies

We included randomized controlled trials which compared different types of PAPR, whether reusable or disposable, for prevention of HCW infection. We included observational studies, defined as studies that follow HCWs over time and that compare the effect of PAPR. We included simulation studies of PAPR technology or alternative respiratory equipment for donning and doffing procedures. In order to maximize study capture, we have chosen a broad range of applicable methodological approaches. Our full eligibility criteria are listed in *Table 5*.

**Table 5.** Review Eligibility Criteria

| *Study characteristic* | *Inclusion criteria* | *Exclusion criteria* |
| --- | --- | --- |
| Types of participants | Healthcare workers<br>Volunteers |  |
| Intervention-treatment | Powered Air-Purifying Respirator (PAPR) studied separately or within a Personal Protective Equipment (PPE). | Hybrid PAPR (hybrid PAPR are designed as both a Self-contained breathing apparatus, PAPR and a standard mask- their design features may not reflect a true PAPR device intended for healthcare use) |
| Comparator | Any other respiratory protective equipment, FFP3/FFP2/N95 or surgical masks. |  |
| Outcomes | -healthcare worker infection rates utilizing PAPR technology within a PPE program as infection with SARS-Cov-2, SARS-Cov-1, EBOLA or MERS;<br>-contamination of skin or clothing measured with any type of test material to visualize contamination;<br>-compliance with guidance on use of PAPR measured with e.g. observation checklist;<br>-level of wearer comfort whilst using the PAPR;<br>-objective and/or subjective measures of work of breathing during the use of PAPR;<br>-costs of resource use of PAPR equipment;<br>-impact of structured training programs on PAPR use; |  |
| Study design | Randomized controlled trials<br>Non-randomized studies<br>Observational studies (cohort studies, case-control studies, cross-sectional studies, case series) | Case reports<br>Surveys |
| Study setting | Inpatient care/Critical Care/Intensive Care; |  |
| Timing | Perioperative process-preadmission, preoperative, intraoperative and postoperative setting | Studies incorporating long-term (greater than three months) postoperative rehabilitation |

### Types of participants

For simulation studies, we included any type of participants (volunteers or HCW) using PAPR or alternative respiratory equipment as part of a protective PPE program. For field studies, we planned to include any HCW exposed to body fluids from patients contaminated with Ebola, MERS, SARS-Cov-1 or SARS-Cov-2.

### Types of interventions

We included studies that evaluated the effectiveness of any type of Purified Airflow Respirator (PAPR), whether disposable or recyclable against suitable face respirators such as N95/FFP2 or any other respiratory protection used. We excluded hybrid PAPR devices due to potential for confounding.

### Types of outcome measures

#### Primary outcomes

We planned to include all eligible studies that have measured:

1. healthcare worker infection rates utilizing PAPR technology within a PPE program for infection with SARS-Cov-2, SARS-Cov-1, EBOLA and MERS;
2. contamination of skin or clothing measured with any type of test material to visualize contamination;
3. compliance with guidance on use of PAPR measured with e.g. observation checklist;

#### Secondary outcomes

We planned to include all eligible measures that have measured the following:

1. level of wearer comfort, visibility and audibility whilst using the PAPR over alternative respiratory protection;
2. objective and/or subjective measures of work of breathing during the use of PAPR versus alternative respiratory protective equipment;
3. costs of resource use including maintenance and cleaning of PAPR equipment;
4. impact of structured training programs on PAPR use over alternative training or no teaching;

### Information sources and literature searches

We searched the following electronic databases: MEDLINE via Ovid SP; EMBASE via Ovid SP; and Cochrane Library (Cochrane Database of Systematic Reviews and CENTRAL). In addition, we sought information from grey literature through the following specific search engines: Google Scholar, OpenGrey, and GreyNet (29-31). We developed a search strategy for Medline via Ovid (*Appendix 1*) and adapted it for other databases. We searched all databases from their inception to the present time. We conducted the original search for studies in May 2020. Due to the dynamic nature of the current pandemic, we repeated our searches in June 2020. We limited our search to English language studies. We did this in order to facilitate the efficiency of the search, bearing in mind that language limitation is unlikely to result in publication bias(32)

### Study Selection

Titles and abstracts of articles returned from initial searches were screened by two reviewer’s *(AL)* and *(AS)* based on the eligibility criteria outlined above. Full texts of potential eligible studies were examined for suitability. References of all considered articles were hand-searched to identify any other potentially eligible studies. Any disagreements were resolved by discussion. The results of the data search were presented in a PRISMA flow diagram indicating the number of studies retrieved, screened and excluded as per exclusion criteria *(see Figure 1*).

**Figure 1.**
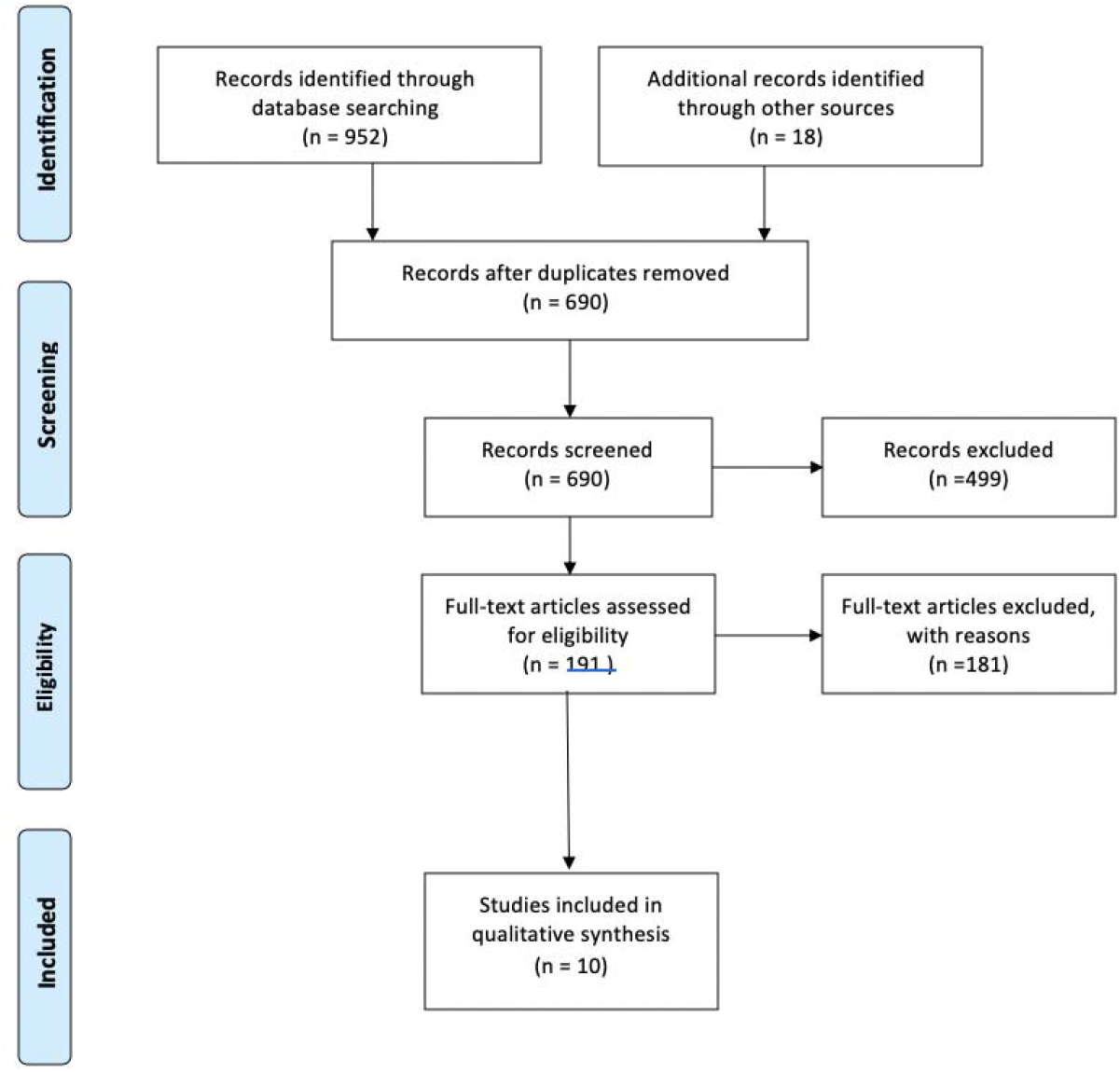
PRISMA Flow Diagram

### Data extraction, management, analysis and presentation

Data were extracted from each study including publication details, study characteristics, participant characteristics, type of procedure, intervention and comparator characteristics, and outcomes. For randomized controlled trials, one author *(AL)* extracted the information on the methodological quality of studies including random sequence generation, allocation concealment, blinding of participants and personnel, blinding of outcome assessment, incomplete outcome data, selective outcome reporting, and other bias (33). For non- randomized studies data were collected on all applicable elements other than random sequence generation and allocation concealment.

### Risk of Bias in Individual Studies

Risk of bias in randomized controlled studies was assessed using the Cochrane Risk of Bias tool (34). We used the ROBINS-I (Risk of Bias in Non-randomized Studies of Interventions) tool to assess the risk of bias in non-randomized studies (35). We rated each potential source of bias as high, low or unclear. We considered blinding separately for different key outcomes where necessary. We used the risk of bias assessment in individual studies to inform our assessment of study limitations across the body of evidence.

### Data Synthesis

We planned to systematically describe the data from each study. We planned to generate Evidence Profile table across each predetermined primary and secondary outcome. We planned to pool data from studies judged to be clinically homogeneous using Review Manager Web Software (36). Due to the heterogeneity of data, quantitative synthesis was not possible.

### Measures of treatment effect

Data ascertained were heterogenous both in terms of study design and interventions undertake. As such, we were unable to estimate treatment effects. We described the included studies in the *‘Characteristics of included studies’* table.

### Confidence in cumulative evidence

Quality of evidence was classified according to the Grading of Recommendations, Assessment, Development and Evaluation (GRADE) system into one of four categories: high, moderate, low, and very low (37). Evidence based on randomized controlled trials was considered as high quality unless confidence in the evidence was decreased due to study limitations, the inconsistency of results, indirectness of evidence, imprecision, and reporting biases. Observational studies were considered low quality; however, they were graded higher if the treatment effect observed was very large or if there was evidence of a dose-response relationship (38, 39). We have presented Evidence Profile (EP) tables in the Appendix section.

## Results

### Results of the search

Our search resulted in 690 references without duplicates for screening (*Prisma diagram, Figure 1)*. Title and abstract screening excluded further 499 studies. We screened the remainder of full text studies. We attained further 18 full text studies through grey literature searches. We included 10 full text studies.

### Included studies

We included ten eligible studies. Please see characteristics of included studies (*Appendix 2*). Five of these studies were simulation studies. Two of the studies were randomized controlled trials. A single study was a randomized controlled trial in a simulation setting (40). A single study was an observational case series of healthcare workers (airway proceduralist only) managing patients infected with SARS-CoV-2 in China at the start of 2020 (41). Two were observational studies with control group cohorts (42) (43). One observational simulation study was a case series without a control group (44).

### Characteristics of participants

In the simulation studies, researchers included 195 participants. Two of the observational simulation studies were cross over studies and therefore control participants were also intervention participants (43) (42, 45-48).

There were 153 participants in the randomized simulation studies. However, 24 of these acted as doffing observers in Andonian et al study (49). There were 1920 on-field healthcare workers performing intubations in two observational studies (41) (50).

### Interventions and comparisons

We identified a large prospective observational cohort study of healthcare workers utilizing a range of respiratory equipment including PAPR devices. The investigators reported that PAPRs (43.4%) were used more commonly in the United States of America (USA) than the United Kingdom (UK). In the UK participants more frequently used FFP3/N100 respirator masks (89.3%). The investigators did not report a significant difference in the primary endpoint rates in these two countries as determined by PPE use (50). We identified a single retrospective observational case series which retrospectively assessed the rates of cross infection in airway proceduralists. In both groups, HCWs utilized droplet precautions with either PAPR (n=50); goggles, FFP2/N95 mask with a face shield (n=22) or goggles, FFP/N95 with a full hood without positive pressure (n=130) (41).

A single randomized controlled trial evaluated the effectiveness of training programs on contamination of personal protective equipment incorporating PAPR (49). A single observational study evaluated attitudes and practices towards a novel PAPR equipment (44). A single observational study compared the effectiveness of different equipment including PAPR on donning and doffing (42). A single observational study evaluated the effectiveness of different respiratory ensembles on temperature of skin and eye dryness (43). A single simulation randomized prospective trial evaluated the PAPR versus E-RCP (40). Three randomized simulation cross over trials evaluated the impact of respiratory equipment including use of PAPR on self-reported wearer comfort measures (45, 47, 48) *Appendix 2* (*Characteristics of included studies*).

### Outcomes

We identified a single prospective observational international multicenter cohort study (El-Boghdadly 2020) reporting the rates of assumed cross-infection with SARS-CoV-2 of healthcare personnel managing the airway. We identified a single observational case series (Yao 2020) which assessed the rates of cross infection in anesthesiologists. In both groups, HCWs utilized droplet precautions with either PAPR (n=50); goggles, FFP2/N95 mask with a face shield (n=22) or goggles, FFP/N95 with a full hood without positive pressure (n=130) (41). We identified no studies assessing the efficacy of PAPR technology compared to alternative respirator/ facepiece during care for patients with SARS-Cov-1, Ebola or MERS;

We identified a single randomized cross-over trial (Zamora et al, 2006) which evaluated contamination of skin or clothing measured with any type of test material to visualize contamination (40); Identified study used fluorescein staining to measure contamination. 26% of participants were contaminated in the PAPR group compared to 96% of contaminated participants in the E-RCP (enhanced respiratory controlled protection) group. We identified a single observational study (42). In a single study (Chughtai et al, 2018) which evaluating the risk of contamination with different PPE and Respiratory Equipment, no participants using PAPR were contaminated. All participants using N95 were contaminated. We found no studies which assessed compliance with guidance on selection of type and use of PPE measured with e.g. observation checklist. We found three observational studies which evaluated the level of wearer comfort, visibility and audibility with using the PAPR over alternative respiratory protection (Chughtai et al, 2018, Chughtai et al 2020, Powell 2017) (42-44).

Three simulation cross-over randomized trials studied the use of PAPR versus APR, with the outcomes of wearer comfort as measured by user rating of mobility, ease of communication, ease of breathing and heat perception (38, 45, 47, 48). We identified no studies which evaluated costs of resource use including maintenance and cleaning of equipment. We identified a single randomized trial which evaluated the utility of training on donning and doffing of personal protective equipment including PAPR (Andonian et al 2019) (49). Structured training using a PAPR decreased the likelihood of self-contamination from 100% to 86%.

### Risk of Bias

We produced a Risk of Bias Summary and a Risk of Bias Graph for individual randomized and observational studies (*Figure 2*) (*Figure 3*). For non-randomized studies (NRS), we identified a high risk of bias across the confounding, selection bias and blinding of outcome assessment across objective and subjective domains. For NRS studies, we identified unclear risk of bias across the blinding performance bias and detection bias, objective outcomes. We used the risk of bias of individual studies to inform our assessment of bias across outcomes. Randomized controlled studies had an unclear risk of bias across a number of domains including allocation concealment, blinding of objective outcome assessment and blinding of participants and personnel.

**Figure 2.**
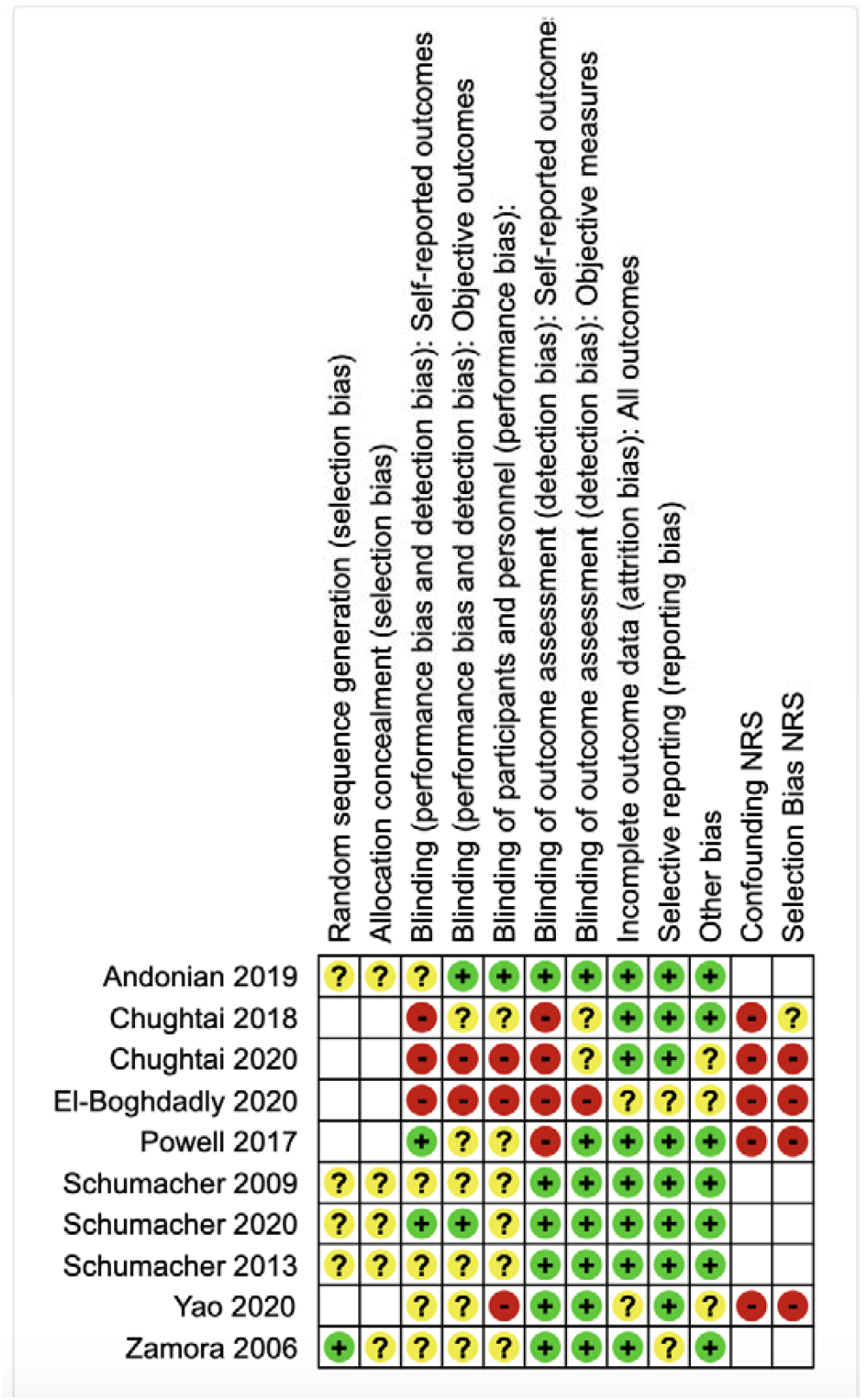
Risk of Bias Summary

**Figure 3.**
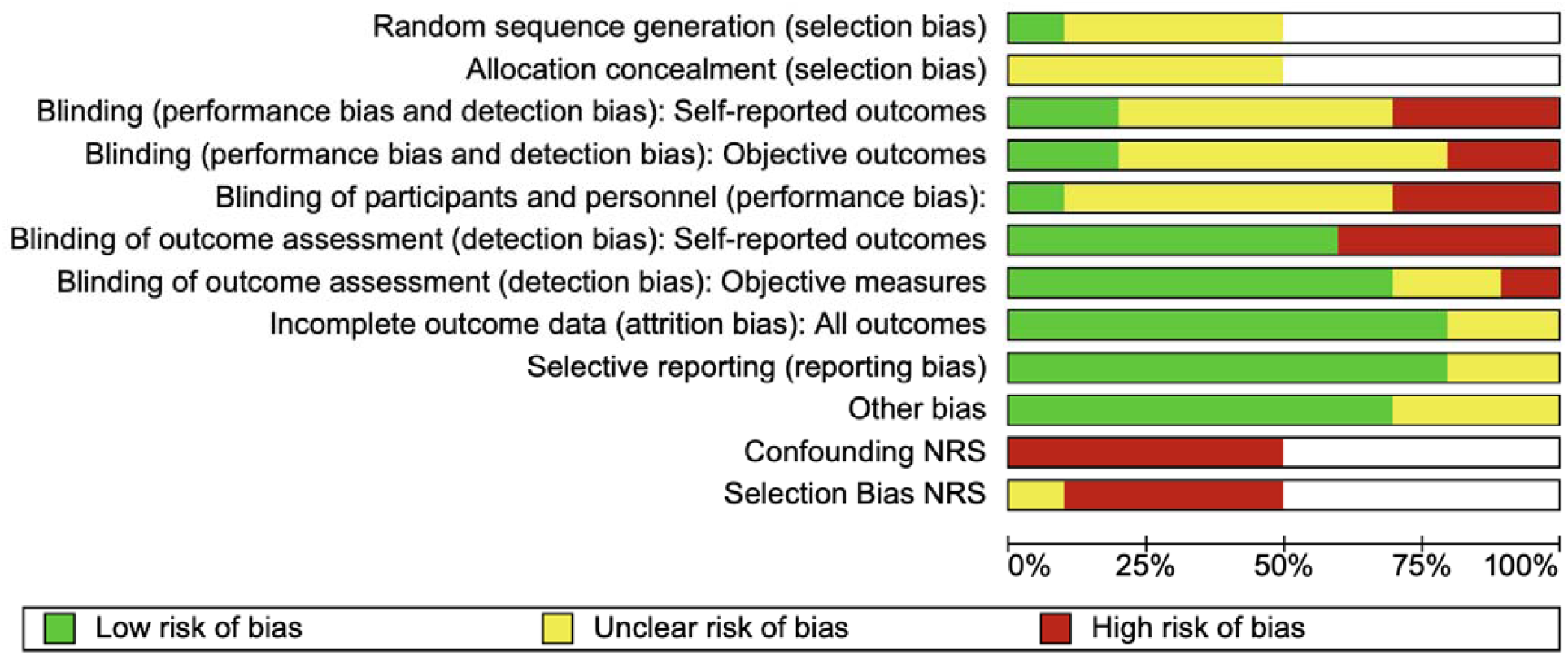
Risk of Bias Graph

### Data Synthesis

We summarized our findings in Evidence Profile *(Appendix 3)* tables across pre-determined primary and secondary outcomes using the GRADEpro software (51). We performed a narrative synthesis of the data.

Data collected were not suitable for a meta-analysis due to inherent heterogeneity. There was no difference in the primary endpoint of COVID-19 infection in respective observational studies in the airway proceduralists utilizing PAPR versus other protective respiratory equipment (50). In the prospective observational study, primary endpoint was defined as incidence of laboratory-confirmed COVID-19 diagnosis or new symptoms requiring self-isolation or hospitalization after a tracheal intubation episode. The overall incidence of the primary endpoint was 10.7% over a median follow-up of 32 days. Most participants were diagnosed through reported symptomatic self-isolation 144 (8.4%). The risk of the primary endpoint varied by country and was higher in females. The risk of COVID outcome was not associated with respiratory protection program use or use of PAPR (50). Investigators did not report the exact number of users protected by PAPR devices. Consequently, we did not construct an EP table for this primary outcome. In the second observational study, there were no airway proceduralists who were cross-infected in either cohort.The rate of healthcare worker infection was significantly different in the two studies, 10.7% versus 0%. Contamination of skin or clothing measured with any type of test material yielded lower risk of contamination in simulation studies. Evidence base for this outcome was low (40, 42).

There was moderate quality of evidence with regards to lower risk of heat build-up in users with PAPR technology (47) (45). There was moderate quality of evidence that visibility was improved in PAPR in comparison with APR (45). There was consistent moderate quality of evidence of decreased user rating of mobility and audibility with the use of PAPR (45, 47, 48). In a single cohort observational study all participants using N95 reported discomfort (42). Powell et al noted a lower temperature measurement in subjects using PAPR (43). This did not translate to a self-reported greater level of comfort in this study.

Participants in a randomized study rated the ease of breathing with the PAPR system significantly better than with the APR (48).

## Discussion

Recently published field studies of HCWs managing patients with COVID-19 demonstrated equivalent rates of healthcare provider infection in cohorts utilizing PAPR versus other appropriate respiratory protection. We identified a trend towards lower level of cross-contamination in participants using PAPR technology compared to alternative respiratory protection in low quality simulation studies. We identified moderate quality of evidence towards improved healthcare worker comfort (heat tolerance and visibility) with PAPR technology compared with alternative respirators. PAPR users scored the technology lower with on mobility, dexterity, audibility and communication. We identified moderate quality of evidence towards improved healthcare worker comfort (audibility and mobility) with APR (airflow powered respirator) technology compared with PAPR.

There appears to be no reported difference in observed infection rates in participants utilizing PAPR or other appropriate respiratory protection. Preferred use of PAPR for respiratory protection may be due to perceived logistical advantages by institutional policy makers. A prospective international multicenter cohort study found no difference in infection rates between cohorts utilizing varied respiratory protection (50). A series published recently found no airway proceduralist infections in the cohort utilizing PAPR versus a cohort equipped with more routine respiratory protection in addition to usual PPE (41). This study was performed retrospectively in Wuhan during the outbreak of SARS-CoV-2. (41). Differences in the airway proceduralist’s COVID outcomes appear distinct: 10.7 % in the El-Boghdadly et al study versus 0% reported in Yao et al (50) (41). These findings may have been confounded by a well-designed enhanced respiratory and contact protective system in the study with no provider infections.

We observed a trend towards lower contamination rates in simulation studies in participants utilizing PAPR (42) (40). These observations are counterintuitive towards an assumption that due to complexity of technology, cross-contamination during doffing with PAPR is more likely. Results of our review demonstrate a trend towards lower HCW contamination rates and decreased doffing violations whilst utilizing PAPR.

We found no studies which assessed compliance with donning or doffing protocols for equipment utilizing PAPR.

In line with subjective reports that PAPR may be more effective in decreasing the effort needed to maintain the work of breathing compared to a more conventional filtering face piece, we identified moderate quality of evidence for this outcome (43) (7). We identified moderate quality of evidence towards improved healthcare worker comfort with regards to heat tolerance and visibility with PAPR technology. It is thought that through the positive air flow, PAPR’s eliminated the heat build up (52). Decrease in audibility and communication difficulties can be anticipated due to increased weight of the equipment and noise generated by positive airflow. In observational studies, we identified a trend toward greater level of self-reported comfort amongst the PAPR wearers (42, 44). Powell et al noted a lower temperature measurement in subjects using PAPR (43). Prior reports have outlined potential for claustrophobia in healthcare providers with field use of PAPR (53). During the tuberculosis outbreaks, use of PAPR’s had a low institutional uptake. This occurred due to a number of factors, including concerns that doctors would appear frightening to their patients and that the motor’s hissing noise would interfere with patient communication (54). Greater acceptance of PAPR by HCWS during both the SARS-Cov-1 pandemic and Ebola may be influenced by HCW perception of relative risk. Khoo et al published a survey illustrating that PAPR as opposed to N95 were more comfortable for HCWs during an Ebola outbreak in Singapore (55).

We identified no studies exploring the costs of resource use of PAPR versus any other filtration pieces. Costs of maintenance of PAPR equipment which require disinfection, cleaning, self-storage, battery maintenance as well as a requirement for education of significant proportion of HCW workforce have not been considered in evidence-based literature. These costs are juxtaposed against more wears per piece of PAPR compared to disposable face-filtering pieces. PAPR use may be a resource utilization prepared strategy for times of greater need for N95/FFP2. It has been noted that there have been fewer equipment shortages for PAPR than N95 (56).

We identified a single simulation randomized controlled trial which demonstrated a trend towards lower risk of contamination when PAPR use was incorporated with a teaching program. During the SARS-CoV-1 outbreak recent training in infection control increased the likelihood of workers adherence to recommended barrier precautions (57). Whilst the initial focus was on use of more stringent respiratory PPE components, further studies found that SARS -CoV-1 transmission was not supported if more standardized PPE was used. Critical system factors protecting the HCWs included compliance with N95 mask application and ongoing use, as well as complementary respiratory protection protocols (25).

Current reports of the choice of protective respiratory technology during the SARS-CoV-2 pandemic are disparate. In a recently published experience of intubation and ventilation of critically ill patients in Wuhan, Meng et al they illustrated use of a positive pressure ventilation system for anaesthesiologists dealing with COVID-19 positive patients (58). There have been three separate descriptive reports from Singapore on routine use of PAPR in their protocols for anaesthesia in suspected or confirmed Covid-19 patients (59) (60) (27). Recommendations from the Joint Task Force of the Chinese Society of Anesthesiology and the Chinese Association of Anesthesiologists center on N95 use for proceduralists. These recommendations do not specifically mention use of a PAPR device. Although some authors make recommendations for the use of PAPR for critical care of Covid-19 patients, they acknowledge that there is no conclusive evidence to show that this advanced respiratory technology decreases the likelihood of viral airborne transmission (61).

Utilization of PAPR with high filtration efficiency may represent an example of “precautionary principle” wherein action taken to reduce risk is guided by logistical advantages of PAPR system. With a higher APF factor than N95 masks, it is scientifically plausible that PAPR use may result in long term lower HCW infection rates. There is however limited literature supporting PAPR use during epidemics/pandemics of SARS-CoV-1, SARS-CoV-2, MERS and Ebola. Given the lack of demonstrable efficacy, institutional decision makers may be applying a pragmatic choice to use PAPR on the basis of precautionary principle.

Current PAPR certification standards have been developed primarily for industrial applications. There is a need for respirator standards to better expand to suit the requirements of healthcare workers (62). In terms of the laboratory research, industrial, radioactive or biological particles behave in a similar manner with regards to a filtration standard. Quantification of the infectious dose with this emerging viral disease has not occurred. Therefore, it remains a challenge to determine the optimum respiratory protection under individual circumstances. Future developments include adjusting the testing standards to activities to which the user (HCW) is engaged.

Our systematic review has been limited by a number of available studies graded as low evidence. A recently published study by El-Boghdadly et al had only 28.8% of laboratory confirmed infections. The remainder were diagnosed through self-isolation and hospitalization without confirmed laboratory testing. (50). In additi in the absence of phylogenetic analysis it is not possible to conclude the source of infection, be it patient contact or community acquired. Comparison of infection rates with HCWs not wearing the PAPR technology may be biased by other PPE protection factors such as the utility of system-related compliance measures (63). Despite the theoretical advantages of PAPR, there have to date been no controlled clinical trials on the efficacy of this technology during the SARS-Cov-1, SARS-Cov-2, EBOLA or MERS pandemics in comparison with other high level respiratory protection(64). At present, minimal infective dose for SARS-CoV-2 pathogen is unknown for any of the transmission modes (65). Higher viral load shedding may be more readily associated with greater disease severity (66). Whether a higher PAPR filtration factor translates to decreased infection rates of HCWs remains to be elucidated. True randomized controlled studies may not be ethically feasible due to higher filtration factor of PAPR. Pragmatic observational studies, as published recently in well-resourced areas may be both more ethical and feasible (41). Most of the studies included have been performed using simulation. Despite simulation being designed to simulate exposure to highly contagious diseases, they are performed in a safe setting without true haste (46). This may introduce systematic bias to the studies themselves and the review. We graded the risk of bias in observational on-field studies as high. This is due to a number of factors including the observational nature of SARS-CoV-2 infection rate assessment and potential for confounding due to attendant infection control processes.

## Conclusion

Equivalent rates of healthcare provider infection have been demonstrated in cohorts utilizing PAPR versus other appropriate respiratory protection. There have been no field studies reporting the effectiveness and utility of PAPR in protecting the healthcare workers from cross-infection due to other highly virulent viral diseases including SARS-CoV-1, Ebola or MERS. Evidence base of low quality indicates greater wearer protection in HCWs using PAPR compared to alternative respiratory devices, from cross-contamination and during doffing in simulation studies. Provider satisfaction appears higher with regards to thermal comfort, however lower in relation to audibility and visibility with PAPR technology. Precautionary principles may be guiding the institutional risk management strategies of HCW protection during epidemics and pandemics.

Closure of this knowledge gap with regards to optimal respiratory protection during pre-defined highly virulent pandemics needs further prospectively collected field data.

## Data Availability

Availability of data and materials
Not applicable

## Abbreviations

(HCW): Healthcare worker
(MERS-CoV): Middle East respiratory syndrome coronavirus
(SARS): Severe Acute Respiratory Syndrome
(PPE): Personal protective equipment
(WHO): World Health Organization
(PAPR): Powered Air-Purifying Respirator

## Declarations

### Author contribution

AL and AS contributed towards the design and conduct of the systematic review, including research questions addressed; RLS contributed towards literature review and analysis of information

### Availability of data and materials

Not applicable

### Ethics approval and consent to participate

Not applicable

### Consent for publication

Not applicable

## Appendices

Appendix 1- Search Strategy;

Appendix 2- Characteristics of Included Studies;

Appendix 3-Evidence Profile Tables;

## Characteristics of Included Studies

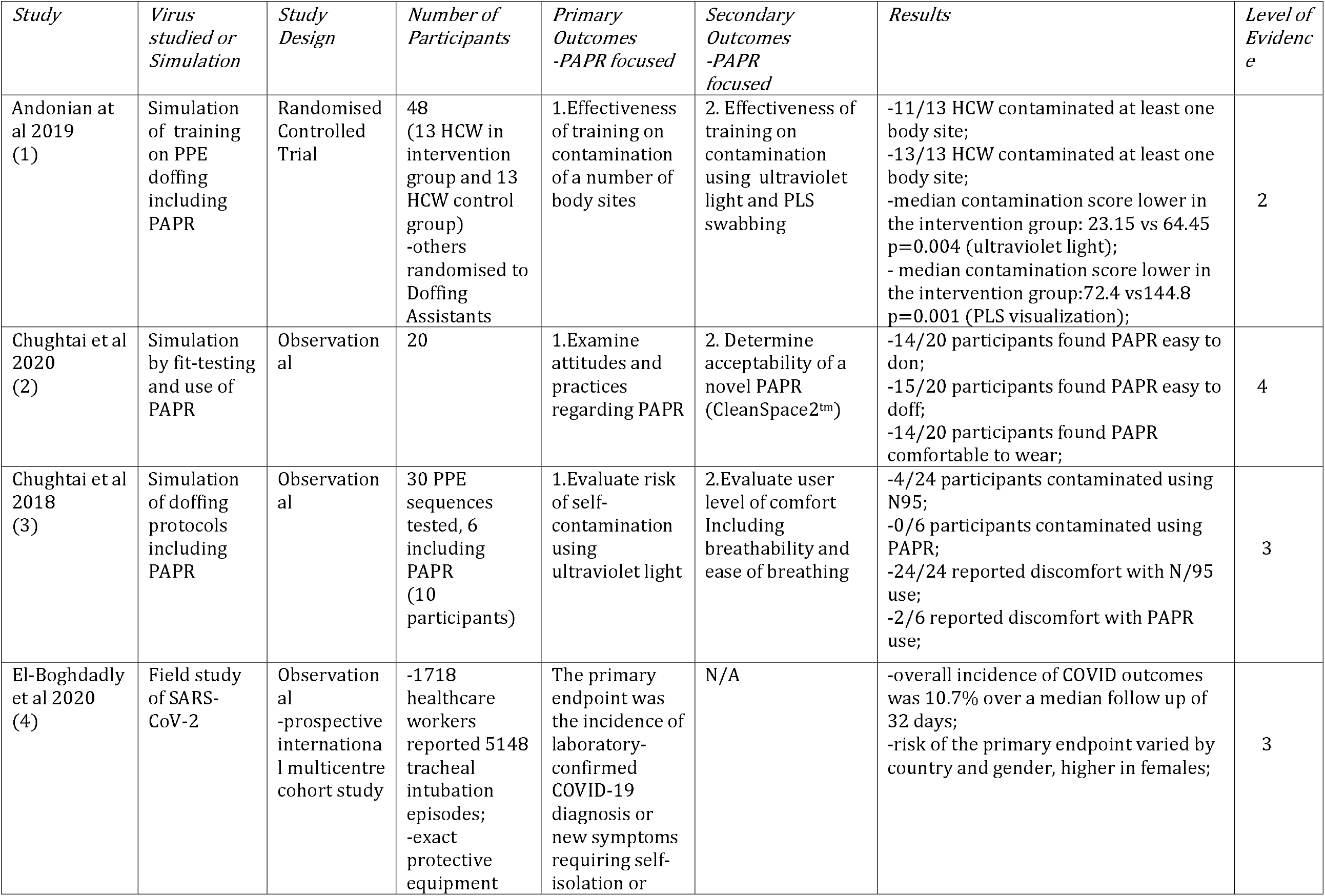
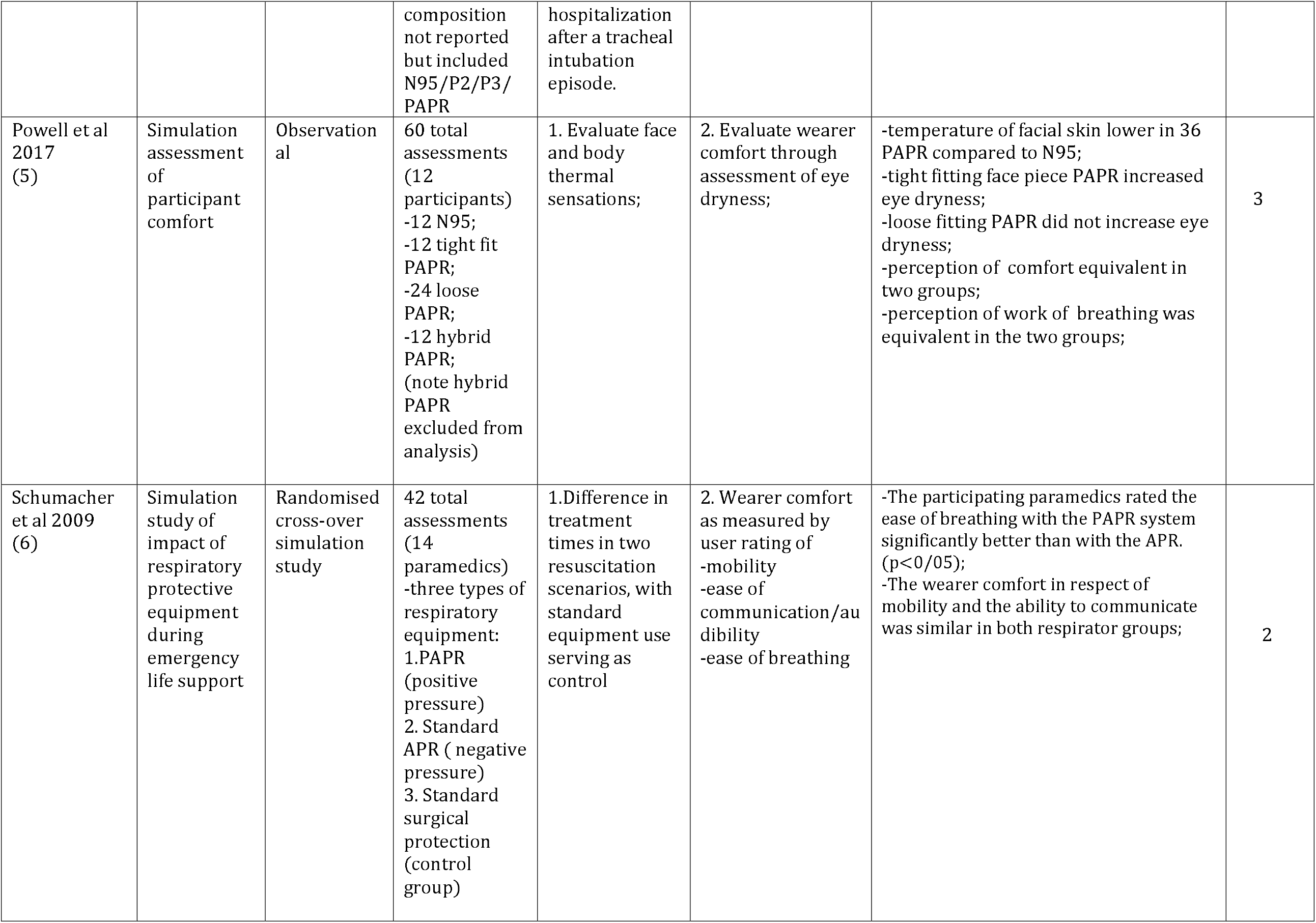
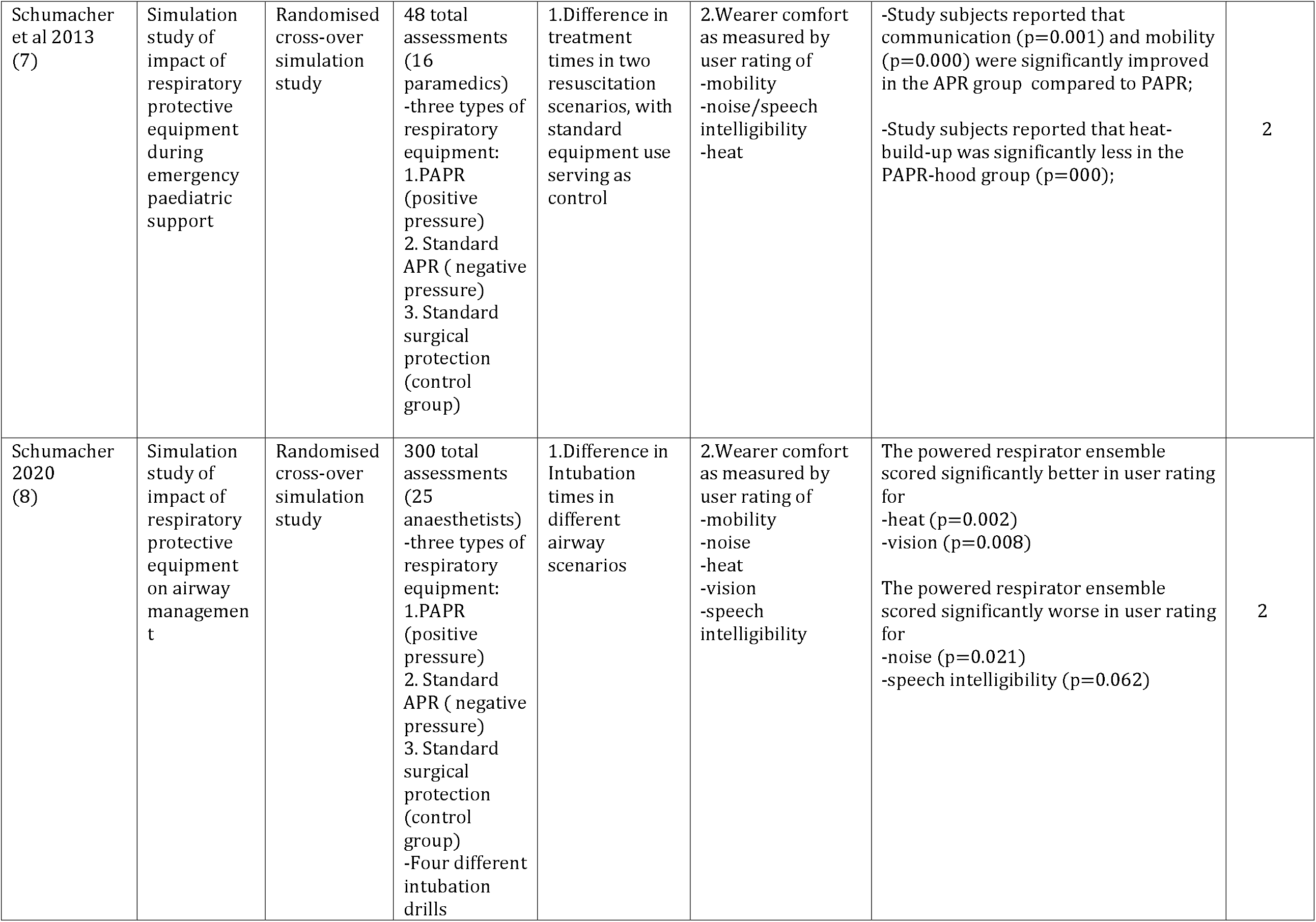
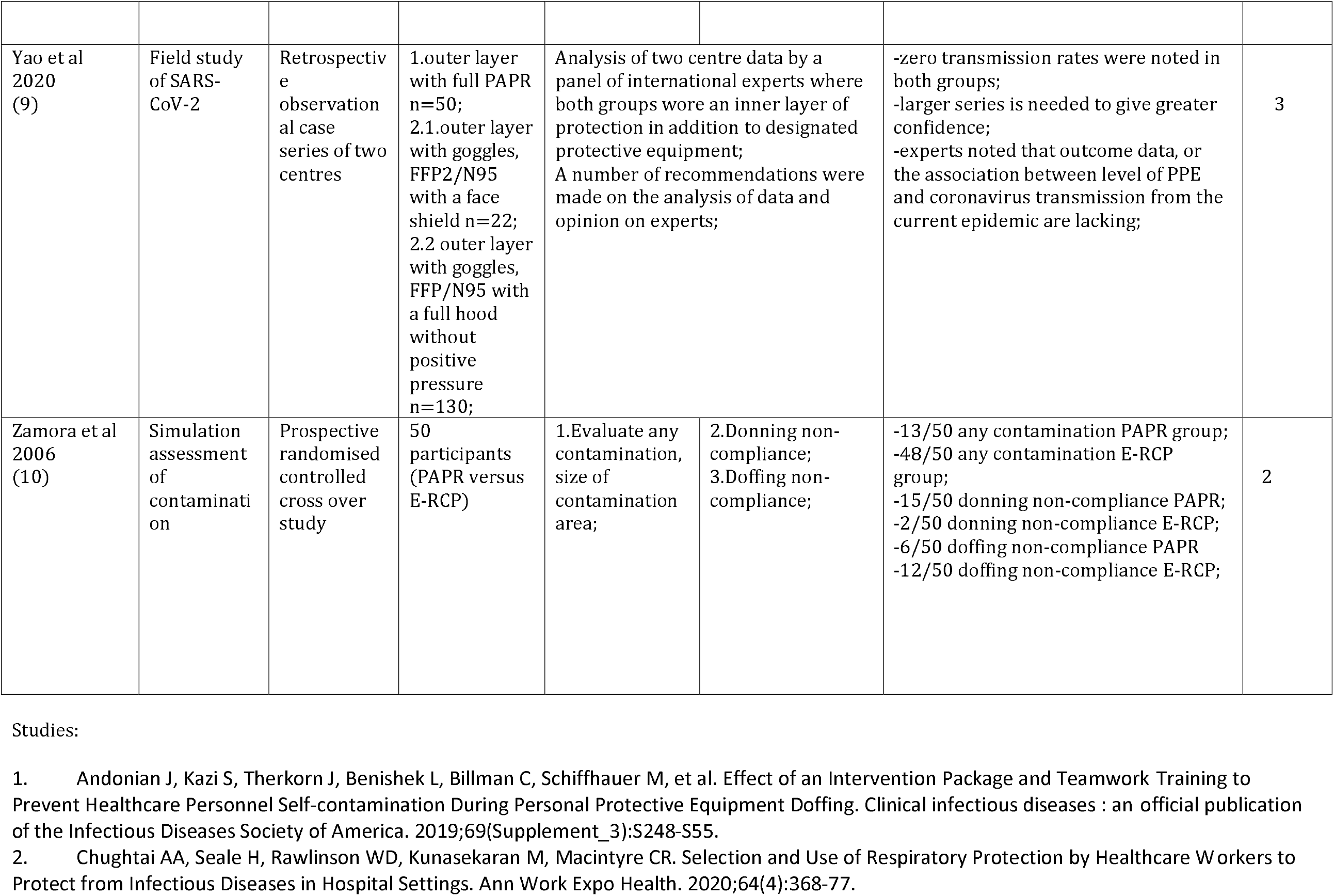
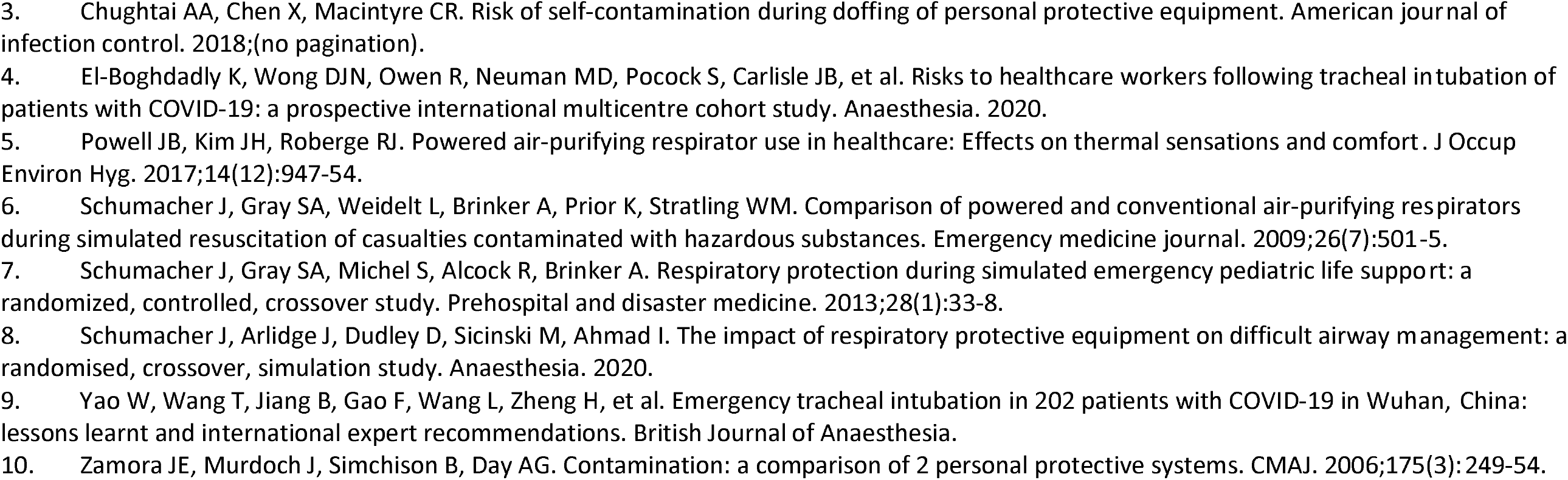

## Primary Outcome: 2

Contamination of skin or clothing measured with any type of test material to visualize contamination;

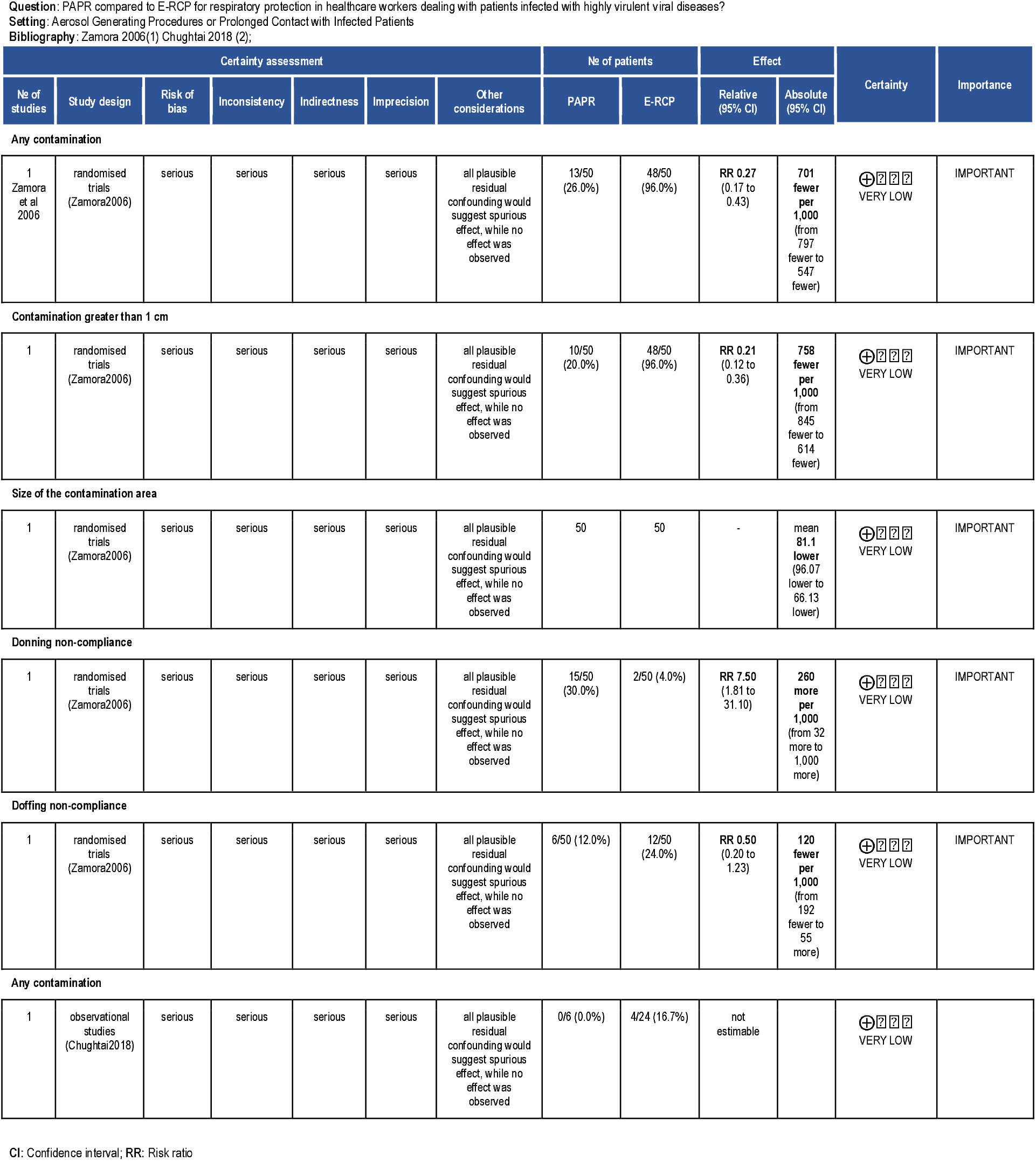

## Secondary Outcomes

1. level of wearer comfort, visibility and audibility whilst using the PAPR over alternative respiratory protection;
2. objective and/or subjective measures of work of breathing during the use of PAPR versus alternative respiratory protective equipment;

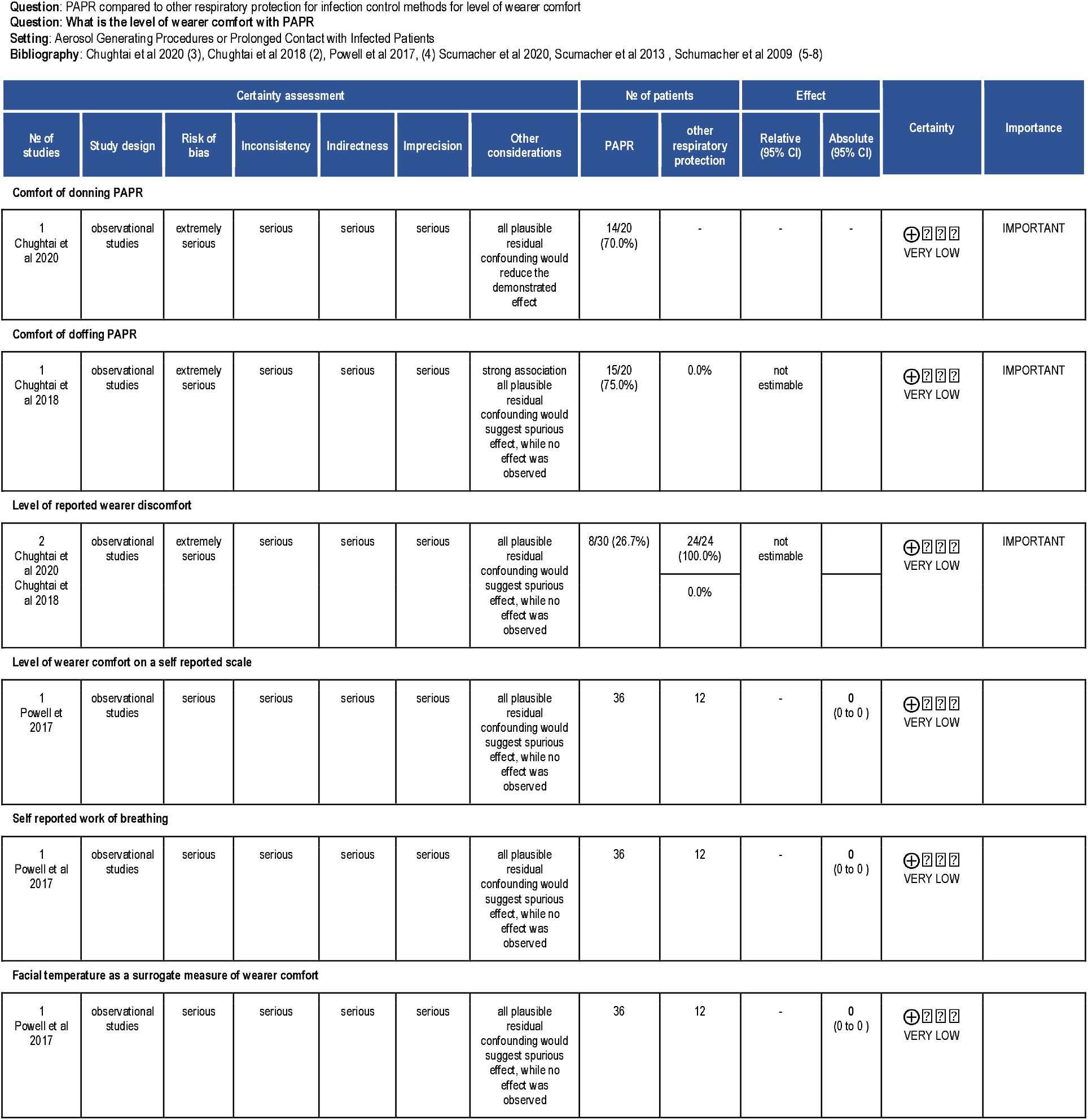
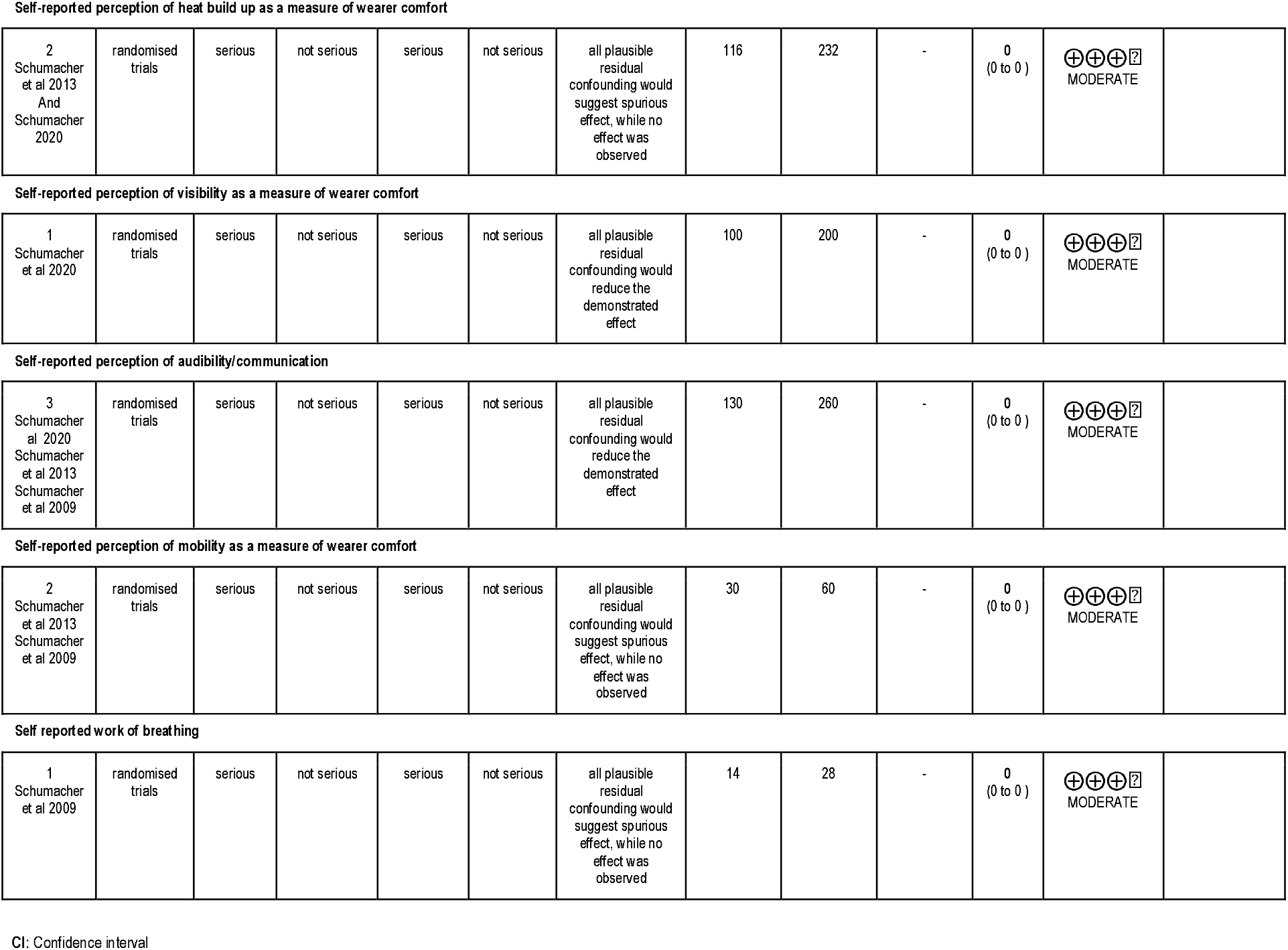

## Secondary Outcome: 4

impact of structured training programs on PAPR use over alternative training or no teaching;

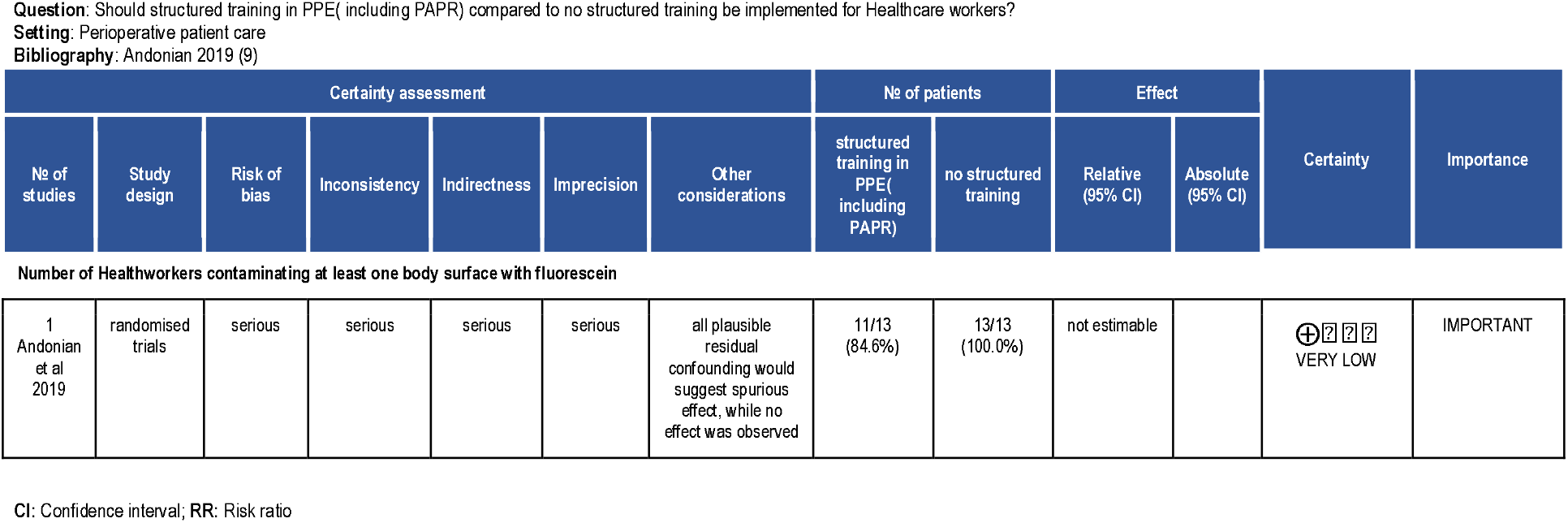

Database: Ovid MEDLINE(R) ALL <1946 to May 12, 2020>

Search Strategy:

--------------------------------------------------------------------------------

1. Respiratory Insufficiency/ or Pneumonia, Viral/ or Lung Diseases/ or Respiratory Protective Devices/ or Pressure Ulcer/ or Continuous Positive Airway Pressure/ or Humans/ or respiratory devices.mp. or Coronavirus Infections/
2. Masks/ or masks.mp.
3. 1 or 2
4. E-RCP.mp.
5. exp Respiratory Protective Devices/ or filtering face piece.mp. or exp Masks/
6. N-95.mp.
7. FFP2.mp.
8. FFP3.mp.
9. air-purifying respirator.mp. or Respiratory Protective Devices/
10. powered air-purifying respirator.mp.
11. 3 or 4 or 5 or 6 or 7 or 8 or 9 or 10
12. exp Health Personnel/
13. nurse.tw.
14. dentist.tw.
15. medical worker.tw.
16. ambulance.tw.
17. physio.tw.
18. physician.tw.
19. transport.tw.
20. 12 or 13 or 14 or 15 or 16 or 17 or 18 or 19
21. 11 and 21
22. Ebolavirus/ or Hemorrhagic Fevers, Viral/ or Hemorrhagic Fever, Ebola/ or ebola.mp.
23. Disease Transmission, Infectious/ or disease transmission.mp. or Communicable Diseases/
24. SARS Virus/ or Betacoronavirus/ or Severe Acute Respiratory Syndrome/
25. Coronavirus Infections/ or Severe Acute Respiratory Syndrome/ or SARS Virus/ or Pneumonia, Viral/ or Betacoronavirus/
26. SARS Virus/ or Pneumonia, Viral/ or Coronavirus Infections/ or SARS-CoV-2.mp. or Betacoronavirus/
27. MERS.mp. or Middle East Respiratory Syndrome Coronavirus/ or Coronavirus/
28. Coronavirus Infections/ or Pneumonia, Viral/ or human-to- human transmission.mp.
29. 22 or 23 or 24 or 25 or 26 or 27 or 28
30. 20 and 29

